# Genomic network analysis links uveitis with systemic inflammatory diseases

**DOI:** 10.64898/2026.03.24.26349228

**Authors:** Karen Chau, Kristen Allison, Tasanee Braithwaite, Isaac T. W. Harley, Lynn M. Hassman

**Affiliations:** Division of Rheumatology, Department of Medicine, School of Medicine, University of Colorado Anschutz, Aurora, CO 80045, USA; Medicine Service, Rheumatology Section, Rocky Mountain Regional Veterans Affairs Medical Center, Aurora, CO 80045, USA; Centre for Translational Medicine, King’s Health Partners, London, UK; School of Immunology and Microbial Science and School of Life course and Population Science, King’s College London, UK; Department of Ophthalmology, School of Medicine, University of Colorado Anschutz, Aurora, CO 80045, USA

## Abstract

**Purpose:** Uveitis is a vision-threatening inflammatory eye disease for which the cause is unknown and response to current therapies is limited. To better understand the genetic architecture of ocular inflammatory disease, we performed a computational synthesis and comparative network analysis of previously published GWAS significant variants associated with 1) uveitis, 2) ocular conditions including age-related and inherited retinal degeneration and 3) extraocular immune-mediated inflammatory diseases (IMIDs).

**Methods:** We identified putative causal genes for genome-wide significance variants from uveitis, IMIDs and ocular diseases using OpenTargets and published studies. To assess the gene-level pleiotropy between disease groups, we quantified the causal gene overlap between groups, and the Jaccard Similarity Indices for individual disease pairs. We then used a network approach to assess the molecular genetic similarity between diseases at a biological pathway level and comparative statistics to identify diseases with greater network similarity to uveitis.

**Results:** Seventy-five percent of the putative causal genes for uveitis were also implicated in IMIDs, while no uveitis genes were shared with primary ocular disorders. Network analysis revealed that 1) uveitis genes are more closely networked with systemic IMIDs disease genes than with ocular-specific disease genes; and 2) significant network similarity links uveitis and specific IMIDs, such as ankylosing spondylitis and sarcoidosis.

**Conclusions:** Overlapping causal genes and network similarity suggest that uveitis is predominantly an inflammatory disease, sharing genetic architecture with other IMIDs.

**Translational Relevance:** These findings provide a genomic framework for understanding the genetic drivers of ocular inflammation in uveitis and generate testable hypothesis for future studies aimed at rationally repurposing drugs from IMIDs for patients with uveitis.

## 1. Introduction

Uveitis, a clinically heterogeneous group of conditions in which inflammation leads to damage of the eye, causes up to 20% of all cases of blindness.^1^ The majority of cases in high-income countries are thought to be immune-mediated and affect 121 per 100,000 people in U.S.^2^ Uveitis is currently treated with anti-inflammatory therapies, however because mechanistic understanding of this disease is lacking, empirically chosen therapies fail to produce disease remission for 30-50% of patients.^3–5^

Genetic analyses of uveitis have implicated several genes that encode proteins with demonstrated roles in immune responses, such as the class I major histocompatibility molecules, HLA-B27 and HLA-A29, as well as the inflammatory cytokine receptor, IL23R.^6–9^ Approximately one-third of uveitis cases are clinically associated with extraocular immune-mediated inflammatory disease (IMIDs), such as ankylosing spondylitis or sarcoidosis,^10,11^ suggesting that common immune mechanisms may drive inflammation across tissues. In agreement with this notion, studies over the past decade have identified a shared genetic architecture across systemic autoimmune diseases, illuminating both common pathways and disease specific mechanisms through a network of interconnected genetic risk variants.^7,8,12^ However, the extent of shared genetic architecture between uveitis and extraocular immunologic disease has not been systematically explored.

Alternatively, given that most cases of uveitis occur in the absence of extraocular manifestations of autoimmunity, eye disease-associated loci may predispose patients to eye-specific inflammation in uveitis. Supporting this notion, inherited structural and degenerative eye diseases result from genetic anomalies that lead to dysfunction of ocular tissues and often present with concomitant ocular inflammation. Likewise, genes predisposing to ocular sarcoidosis are implicated in both barrier function as well as autoimmunity.^12^

Resolving the genetic architecture of uveitis is critical for identifying both new therapeutic targets and opportunities for drug repurposing opportunities to benefit the 30-50% of patients for whom empiric treatments fail. To date, the genetic association between uveitis and both ocular disorders and systemic IMIDs has not been systematically explored. To better resolve the pathophysiologic similarities between uveitis and both eye-specific and immune-mediated diseases, we performed a hypothesis-generating computational synthesis of published GWAS data using gene-level overlap and inferred biologic networks. This synopsis suggests testable hypotheses that may yield new therapeutic targets, a rational approach to drug repurposing, and the potential for endotype-stratified clinical trials for patients with blinding ocular inflammatory disease.

## 2. Materials and Methods

### Sources of genetic data for disease phenotypes

The GWAS data for uveitis in our analysis was derived from 25,377 non-infectious uveitis cases (24,294 with anterior uveitis/iritis, 774 with Vogt-Koyanagi-Harada disease, 192 with uveitis associated with juvenile idiopathic arthritis (JIA) and 117 with birdshot chorioretinopathy) (Supplemental Table 1). We also included hereditary ocular diseases (i.e. structural eye diseases, juvenile glaucoma and retinal dystrophies), as well as age-related macular degeneration (AMD), and genes associated with optical coherence tomography (OCT).^13^ We included monogenic inflammatory diseases (i.e. inborn errors of immunity such as Blau Syndrome, Familial Mediterranean Fever) as well as polygenic IMIDs such as Juvenile Idiopathic Arthritis and Lupus. We included osteoarthritis as a control.

### Causal Gene Identification

We identified single nucleotide polymorphisms (SNPs) associated with uveitis and each inflammatory or ocular disease in our study from the following sources: National Human Genome Research Institute – European Bioinformatics Institute (NHGRI-EBI) GWAS Catalog, InFevers (online database of genes associated with autoinflammatory disorders), Genomics England panels, the Ocular Knowledge portal, and published papers (Supplemental Table 2). We then identified putative causal genes for each variant of genome wide significance (p<5E-8) using the Open Targets Genetics platform^14^ (Table 1 and Supplemental Table 1). The Open Targets Genetics platform is a collaborative initiative hosted by a consortium of organizations including the Wellcome Sanger Institute, EMBL European Bioinformatics Institute, Genentech, GSK, MSD, and Pfizer. For studies available on the site, we used the Locus-to-Gene (L2G) score, a disease-specific approach that integrates fine-mapping, functional genomics data, and machine learning. The L2G score is calibrated so that a higher score indicates a higher probability that the gene is causally associated with the disease.^15^ For papers not yet integrated into Open Targets Genetics, we used the older Variant-to-Gene (V2G) score, which is a more general, disease-agnostic approach. The V2G score aggregates data from various datasets such as chromatin interaction, quantitative trait loci (QTLs), and in-silico functional predictions, but treats variants as independent.^16^

**Table 1.**
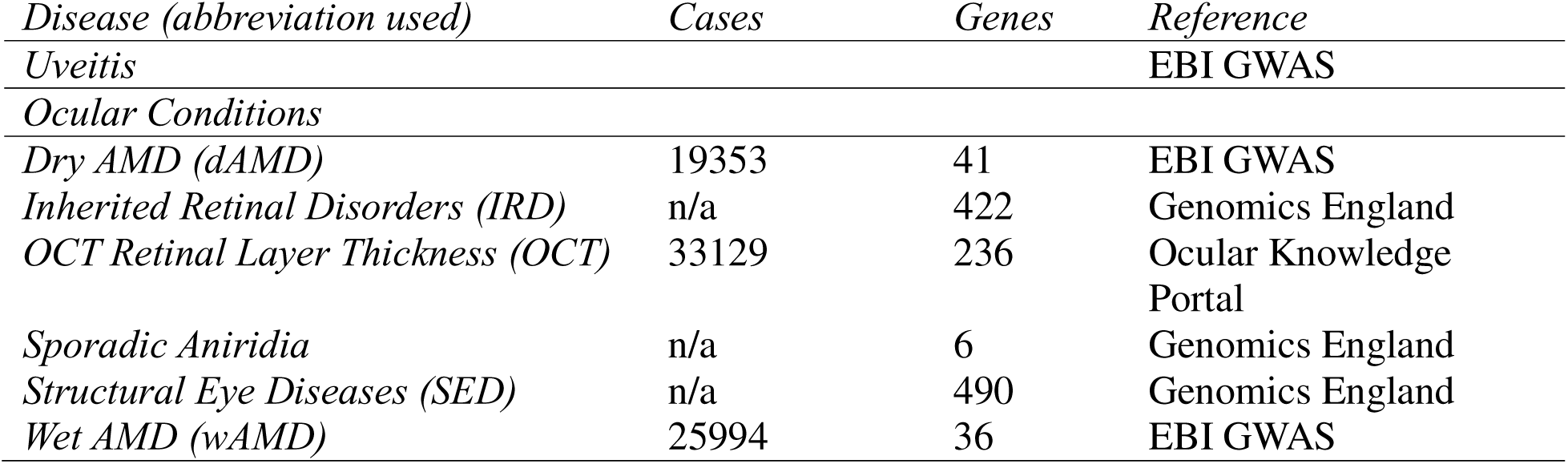
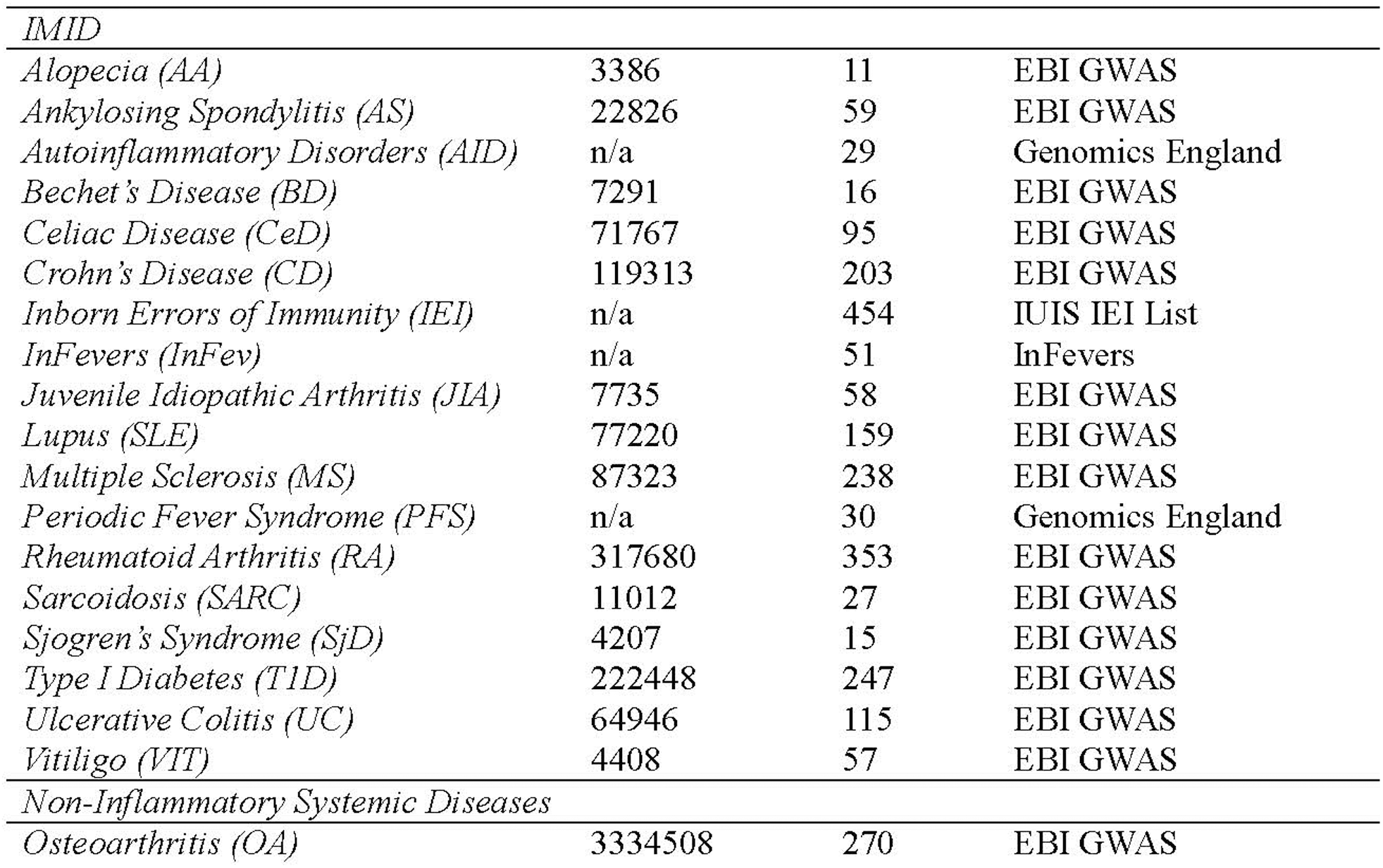
Diseases included in this analysis. The number of cases available for each disease from GWAS published to date, the number of associated genes reaching genome wide significance level of 5×10-8, and source references. *Full gene lists and notes can be found in Supplemental Table 4 and 5*.

### Network Analysis

We loaded the putative causal gene list of L2G and V2G genes for each disease and disease pair into the free version of STRING database for network analysis, generating gene networks displaying the genes as nodes, and predicted protein-protein interactions as edges.^17^ Proteins were searched in the Homo Sapiens database with evidence based edges and medium confidence (STRING score of 0.4) for the interactions. We chose the text mining, experiments, databases, co-expression, neighborhood interactions, gene fusion and co-occurrence as our active interaction sources. The networks were subsequently exported for further analysis and clustering in Cytoscape, an open source platform for complex network analysis^18^ and displayed using the perfused force-directed layout, which positions nodes with stronger interactions closer together.^19^ That is, shorter edges have a higher combined channel information score as calculated in the STRING database.^20,21^

For each two-disease comparison, the predicted protein-protein interaction ratio was defined as the number of direct interactions (edges) between genes from uveitis and the targeted condition, divided by the total edges in the network of the targeted condition and uveitis. The PPI enrichment values quantify the degree to which the protein interactions within a specific set are more significant when compared to a random network of genes. A smaller PPI enrichment suggest that the proteins are highly interconnected, forming a cohesive network that is likely to represent a shared biological function or pathway, thus enabling a more focused and meaningful analysis.^22^

Some genes may not appear as nodes because they are not represented in the STRING database. STRING integrates data from genomic context predictions, high-throughput experimental studies, co-expression analyses, automated text mining, and curated information from existing databases to construct its comprehensive network of protein-protein interactions.^23^ Genes that are less well characterized in the literature or experimental datasets are less likely to be included. For example, in the context of uveitis, 19 out of 20 genes were represented as nodes, whereas HLA-B27 was combined with HLA-B with the STRING visualization.

The Jaccard similarity index is a metric used to quantify the similarity between two sets by comparing the number of shared elements to the total number of unique elements across both sets. In this study, we applied it to measure the overlap of genes associated with different diseases. Specifically, for each disease, we compiled a list of associated genes and then computed pairwise comparisons between these gene sets. The Jaccard index J(A,B) for two sets A and B is defined as 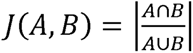, where *A ∩ B* s is the number of shared genes between the two disease and *A ∪ B* is the total number of unique genes across both diseases. A higher Jaccard index indicates a greater degree of genetic overlap.

### Statistical Analyses

To assess the significance of shared edges (protein-protein interactions) between uveitis-associated genes and other diseases, we applied a hypergeometric test. Let N denote the total number of edges in the human interactome, the set of all human protein to protein interactions, or all known human proteins contained in STRING; M the number of edges within one degree of a given disease; n the number of edges within one degree of uveitis genes, and k the number of edges observed in common between the two. The probability of observing k or more shared edges by change is given by the hypergeometric survival function:

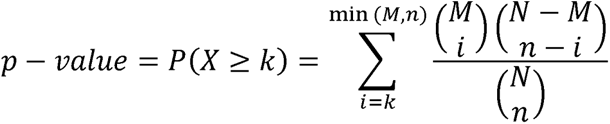

A lower p-value indicates a greater than expected overlap, suggesting a significant network-based relationship between the disease and uveitis genes.

For the analysis in Figure 4, we define shortest path as follows. Shortest path is the minimum number of edges between a node or subset of nodes in the network and another node. As applied to Figure 4, nodes denoted as one degree separated (1°) are within one edge of the subset of nodes representing the Uveitis genes. Nodes denoted as two degrees separated (2°) are at least two edges separated from the nodes representing the Uveitis genes. Similarly, nodes denoted as three degrees separated (3°) are at least three edges separated from the nodes representing the Uveitis genes. Finally, nodes denoted as (>3°) are at least four or more edges separated from the nodes representing the Uveitis genes.

Chi-squared analysis of this network evaluated the association with disease/phenotype category and degree of separation from uveitis. Specifically, a 2X4 table was constructed. The rows consisted of IMID or eye phenotype genes (categories as defined in Figure 4 legend). The columns consisted of Degree of separation from the subset of uveitis genes. Standard Chi-squared analysis was carried out using Microsoft Excel as detailed in Supplementary Table 3.

## 3. Results

To assess the extent of genetic overlap between uveitis and either eye-specific developmental and degenerative diseases or immune-mediated inflammatory diseases (IMIDs), we identified putative causal genes from published SNPs of genome wide (GW) significance for uveitis, inherited, degenerative and structural eye diseases, inflammatory and autoimmune diseases, and osteoarthritis, a disease that shares end organ damage, but not immunogenetic risk factors, with several IMIDs. We also included monogenic causes of inherited ocular and systemic inflammatory diseases (Table 1).

### Uveitis shares more predicted causal genes with inflammatory diseases than other eye diseases

The GWAS data in our analysis was derived from 25,377 non-infectious uveitis cases of which 95% represented anterior uveitis. In contrast, many of the extraocular IMIDs we included have been more widely studied using GWAS, for example, Rheumatoid Arthritis and Multiple Sclerosis included approximately 317,680 and 87,323 cases, respectively. Thus, the opportunity for identification of shared putative causal genes is somewhat limited by the lower number of studies in uveitis. Despite this, we found that 75% (15/20) of putative causal uveitis genes were also associated with extraocular IMIDs (Figure 1). The uveitis genes overlapping with IMIDs included *ADO, B3GNT2, C1orf141, EGR2, ERAP1, HLA-A, HLA-B27, IL23R, MICA, HLA-DRB1, LNPEP, HLA-DQA1, HLA-B, LYRM9,* and *TNRC18. CAVIN4, DPP10, HS3ST5, OR2L13,* and *SELENOV* did not overlap with the IMIDs.

**Figure 1.**
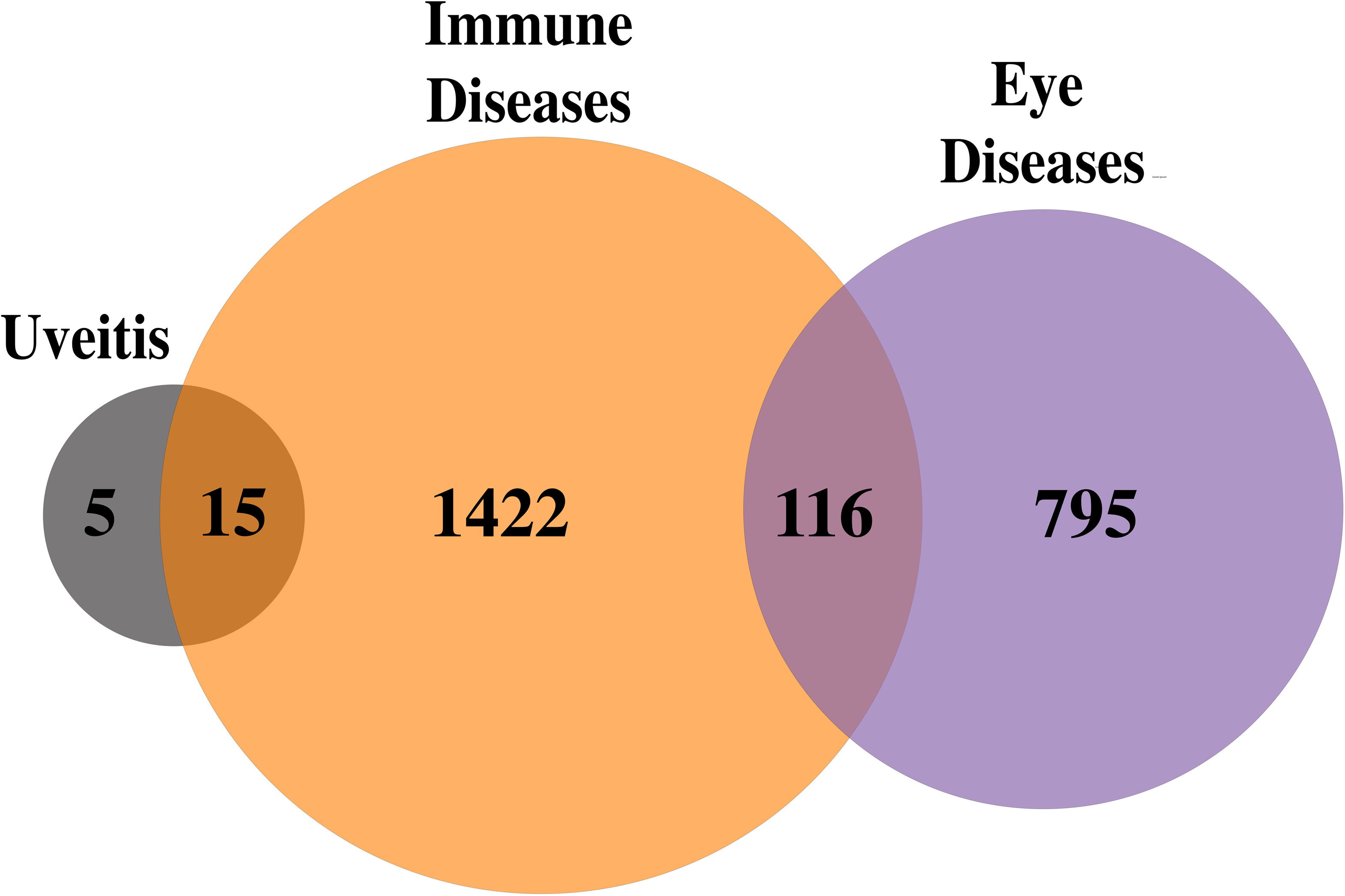
Predicted causal genes overlap between uveitis and Inflammatory Diseases. Venn diagram showing 15/20 putative causal genes for Uveitis are shared with Immune-mediated Inflammatory Diseases and no uveitis genes are shared with non-uveitis Eye diseases (*list of diseases and abbreviations in Table 1*).

As most non-infectious uveitis cases occur in the absence of extraocular inflammatory disease, we also asked whether uveitis shares genetic similarity with inherited or degenerative eye diseases. In contrast to the significant overlap with systemic autoimmunity, there was no overlap between uveitis and the other ocular disorders in our study, despite the presence of shared causal variants between non-uveitis ocular diseases and systemic autoimmunity.

### Common causal genes drive uveitis and a subset of inflammatory diseases

Next, to identify IMIDs with the greatest genetic overlap, we assessed the proportion of shared putative causal genes between each disease in our cohort as Jaccard similarity (Figure 2). We identified several expected associations, including a high degree of overlap between diseases that affect the same target organ, such as Crohn’s disease and Ulcerative Colitis, and between dry (non-neovascular) and wet (neovascular) age-related macular degeneration (AMD). We also observed a high degree of overlap between co-morbid diseases, chiefly between type I diabetes (T1D) and rheumatoid arthritis (RA), consistent with the 5-fold increased risk for RA patients with T1D.^24^

**Figure 2.**
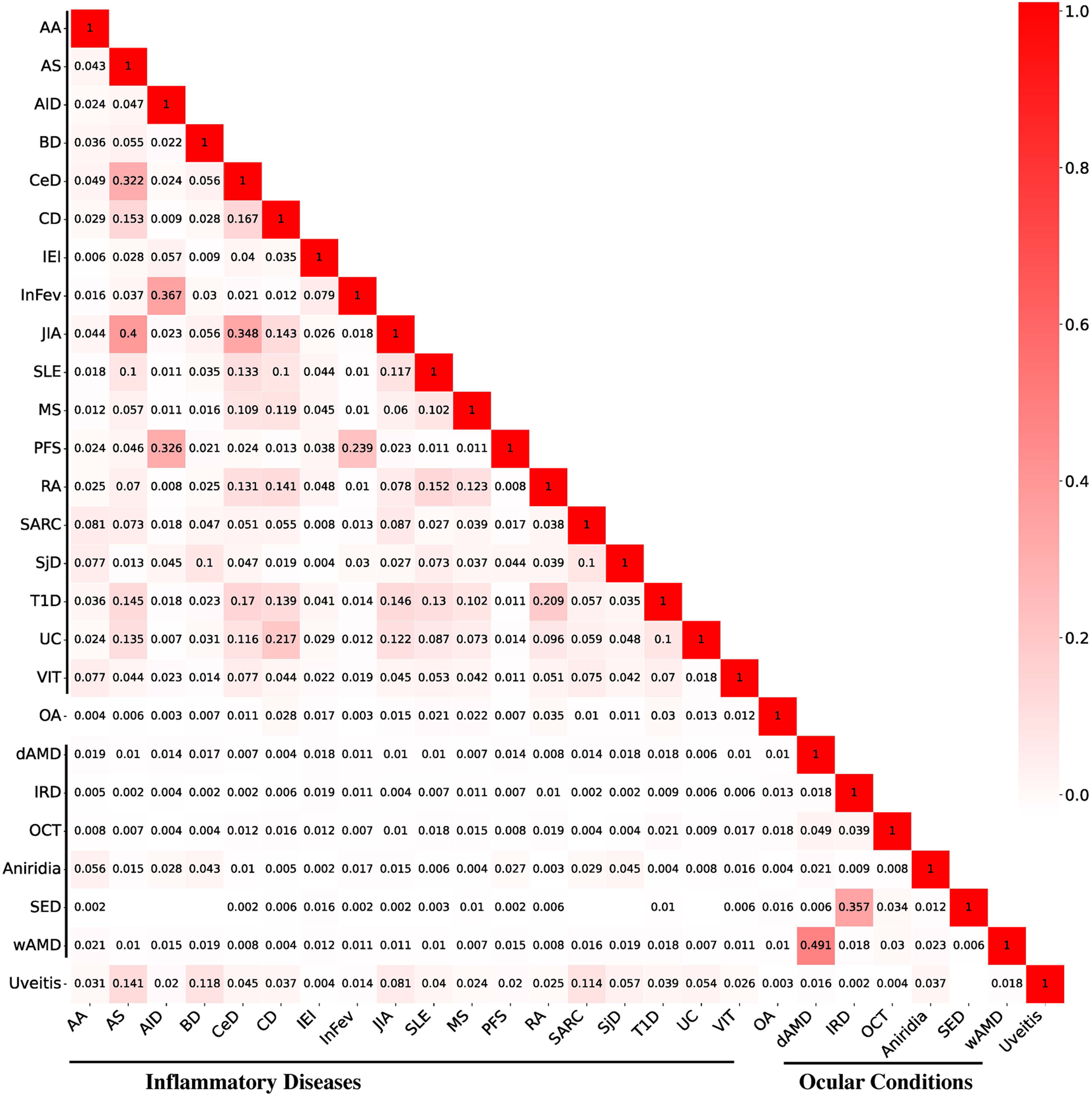
Common causal genes drive uveitis and a subset of inflammatory diseases. Heatmap of Jaccard Similarity Indices for the pairwise genetic overlap between each disease based on available published GWAS studies.

Extraocular inflammatory diseases clinically associated with uveitis, including ankylosing spondylitis, sarcoidosis, Behcet’s disease, and juvenile idiopathic arthritis, had the strongest genetic overlap with uveitis. The similarity between uveitis and individual IMIDs, however, was less strong compared to the similarity between T1D and RA, diseases that were significantly overrepresented in our analysis.

In agreement with previous findings, we did not detect significant genetic overlap between a non-autoimmune joint disease, osteoarthritis, and the inflammatory diseases we studied, including uveitis.^25,26^ Likewise, we did not detect an overlap between uveitis and inherited retinal disorders, structural eye diseases, wet or dry AMD.

### Causal genes in uveitis and clinically associated inflammatory diseases share network relatedness

Next, we constructed predicted protein-to-protein interaction networks utilizing STRING database^17^ using the putative causal genes for each disease. Uveitis genes alone exhibited more interaction than would be expected by chance (PPI enrichment value of P < 1.63E-11). From 19 genes, we identified 14 interactions with close interaction between 8 genes (Figure 3A, Supplemental Table 4).

**Figure 3.**
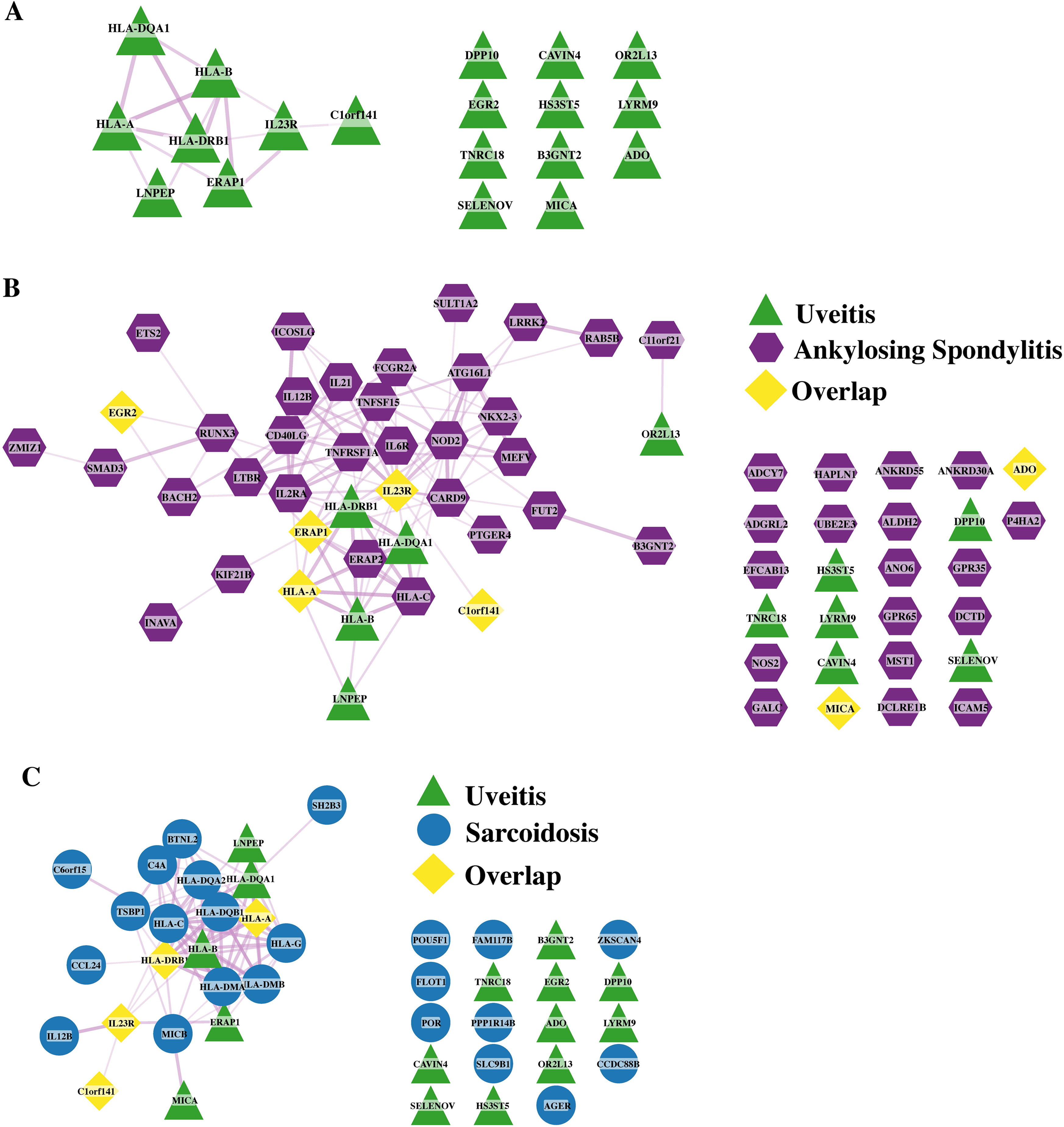
Causal gene relationships in uveitis and clinically-associated inflammatory diseases share network relatedness. **(A)** The predicted protein-protein interaction (PPI) STRING network from GWAS-identified genes for uveitis **(B)** The PPI STRING network from GWAS-identified genes for uveitis and ankylosing spondylitis (AS). **(C)** The PPI STRING network from GWAS-identified genes for uveitis and sarcoidosis. Green represents uveitis, purple represents AS, blue circles represent sarcoidosis, and yellow represents the overlap between the uveitis and AS (B) or sarcoidosis (C).

We then assessed the network relationship between uveitis and individual IMIDs. Uveitis occurs in 33-40% of patients with AS, ^8,27^ making these two conditions the most comorbid in our analysis. Consistent with this clinical comorbidity, we found a high degree of network similarity between ankylosing spondylitis (AS) and uveitis, including 9 overlapping putative causal genes, 7 of which were linked via STRING networks (Figure 3B). Notably, the number of network-connected uveitis genes was greater when uveitis and AS were analyzed together than when uveitis was analyzed independently, consistent with the hypothesis that clinically related diseases share a genetic architecture of biologically related loci.

The uveitis-AS overlapping genes converged around three biologically coherent axes: innate immune activation (*NOD2*, *CARD9*, *MEFV*, *TNFRSF1A*), antigen presentation to T cells via *ERAP1* and MHC class I, and T cell costimulatory molecules (*IL23R*, *IL12B*, *CD40LG*, *ICOSLG*). The strong clinical overlap and specificity of genetic overlap suggest that AS-related uveitis may represent a discrete pathophysiologic endotype within the broader category of ocular inflammatory disease. Furthermore, several therapeutically targetable pathways are implicated by the strongest overlapping genes between uveitis and AS: *IL23R*, for which FDA approved therapies exist, and *ERAP1*, for which therapies are in clinical development. Together, this provides a rationale for endotype-stratified clinical trials targeting the IL-23 pathways or inhibiting ERAP for patients with AS-related uveitis.

Uveitis affects 5-30% of patients with sarcoidosis, so we next applied STRING network analysis to a combined analysis of uveitis and sarcoidosis, using 27 putative causal genes for sarcoidosis and 19 genes for uveitis.^28–31^ Four putative causal genes, including 2 major histocompatibility (MHC) class II loci, *IL23R* and a proximal open reading frame *C1ORF141*, were directly shared between uveitis and sarcoidosis.^28–30^ Additionally, the number of STRING network-linked genes (edges) increased from 14 in the uveitis alone network to 85 in the combined uveitis-sarcoidosis network (Figure 3 and Supplemental Table 4), indicating a shared genetic architecture between sarcoidosis and uveitis, consistent with their high degree of comorbidity.

The uveitis-sarcoidosis network centered around both MHC class I and II alleles, consistent with prominent T cells in biopsy samples from each disease. There was additional overlap around *MICA/B*, suggesting a role for NK cells in each disease. Finally, like AS and uveitis, sarcoidosis and uveitis shared risk alleles in *IL23R* and *IL12B* (part of the IL-23 dimer), further highlighting the therapeutic potential for targeting the IL-23 pathway in uveitis, and potentially expanding the subset of patients who might respond to IL-23 targeted therapy beyond those with AS-associated uveitis.

### Genetic risk for uveitis and inflammatory diseases is related across biologic networks

Next, we systematically assessed the biologic overlap between uveitis and other inflammatory and ocular conditions. We considered the number of shared genes (gene overlap), the proportion of shared edges (i.e. the number of shared edges in each uveitis-single disease network divided by the total number of edges in an all-disease combined network), and the hypergeometric p-value, which indicates a greater than expected overlap between genes from each disease and uveitis (Figure 4).

**Figure 4.**
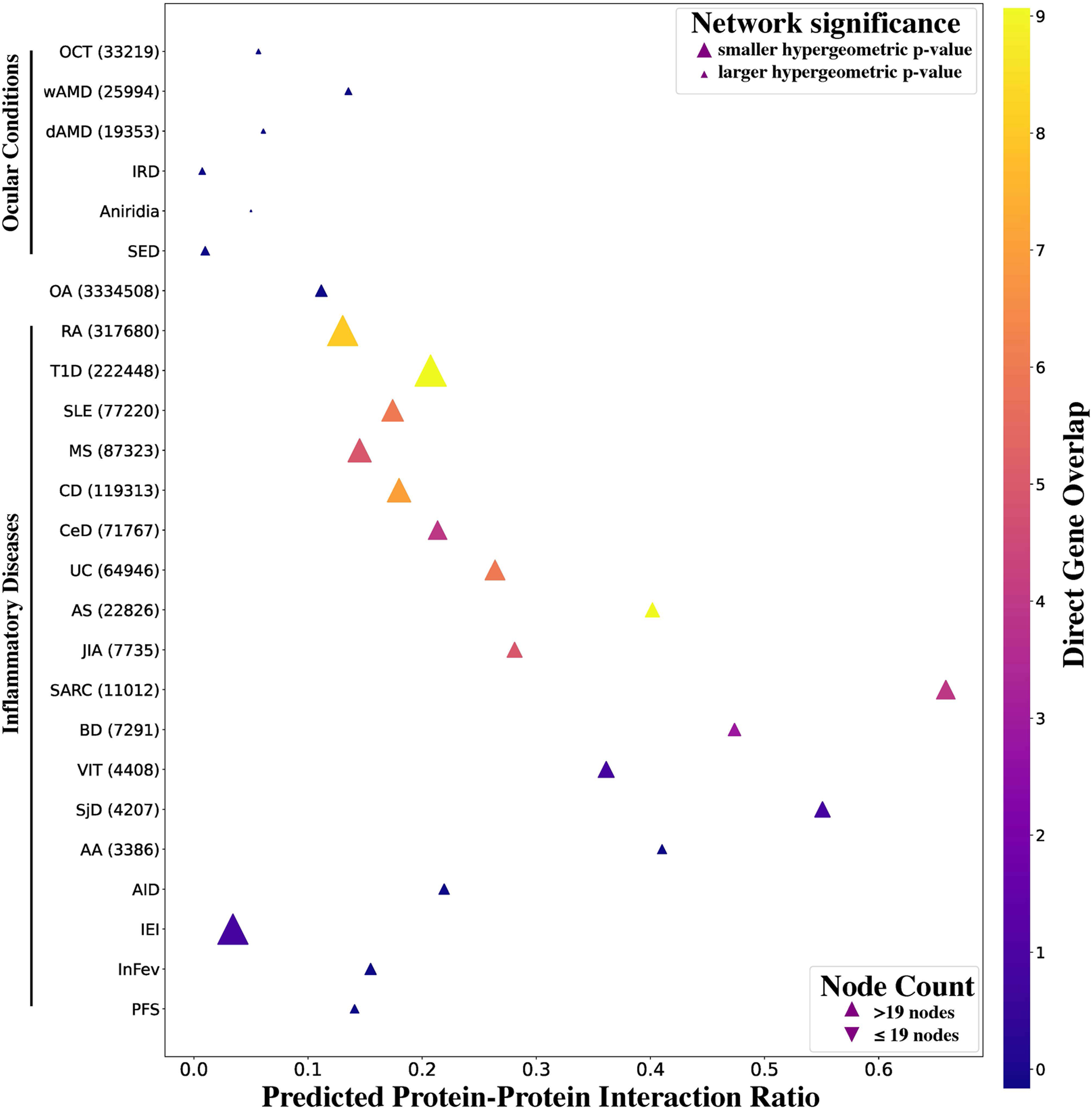
Genetic risk for autoimmune disease including uveitis is related across biologic networks. Predicted protein-protein interaction networks were constructed between uveitis and each disease represented on the y-axis. The number of directly overlapping genes between uveitis and the other conditions in our cohort is represented by triangle color (right legend). Predicted protein to protein interaction ratio (x-axis) is the quotient of the edges that exist between each condition’s network and the uveitis network over the total edges in a network constructed from both diseases together. The number of nodes in the combined network is indicated by direction of triangle (up: >19, down: ≤19). Network significance, i.e. the predicted protein-protein interactions between uveitis and the disease of interest vs. the total human gene database is indicated by triangle size (increasing triangle size indicates smaller hypergeometric p value).

Only the conditions classified as immune-mediated or inflammatory exhibited significant combinatorial overlap with uveitis. Of these, the largest data sets, T1D and ankylosing spondylitis (AS) had the highest direct overlap with uveitis, both sharing 9 genes, followed by Rheumatoid Arthritis (RA) and Crohn’s disease (Figure 4 and Supplemental Table 4). Unsurprisingly, networks comprised of uveitis and these well-studied IMIDs had small hypergeometric p-values indicating a high number of significant protein-protein interactions. Inborn errors of immunity also had a small hypergeometric p value. Two important differences between the individual IMID lists and the IEI list warrant consideration in interpreting this result. First, the IEI network is large (456 genes) in comparison to some of the other diseases. This could inflate the relative significance of association in comparison with polygenic diseases like T1D and RA. Second, the IEI list represents a phenotypically diverse group of monogenic diseases, which may be connected via an incompletely defined polygenic network of immunomodulatory genes. In support of this notion, we previously found that monogenic and polygenic forms of systemic lupus erythematosus (SLE) genes are highly connected in the STRING network and thus likely inhabit the same immune signaling pathways.^32^

Notably, despite the relatively small data set (11,012 cases, 27 putative causal genes), sarcoidosis had a relatively high overlapping node count with uveitis, as well as the highest predicted PPI and a significant hypergeometric p value. This suggests that uveitis may share more genetic architecture with sarcoidosis than previously appreciated. Similarly, Sjogren’s syndrome, represented by a small number of cases (4207 cases, 15 putative causal genes) had a high predicted PPI with uveitis, suggesting that while, underrepresented in our analysis, may have an outsized genetic similarity with uveitis.

### Uveitis is more closely related across biologic networks to immune-mediated diseases than to other ocular diseases

Finally, we sought to determine whether putative causal genes from IMIDs were more likely than eye disease-related genes to directly interact with putative causal uveitis genes. To do so, we analyzed a network that included all the uveitis genes (Figure 3), as well as genes that were only implicated in a single phenotype in addition to Uveitis (Figure 5). This single phenotype network included 2312 genes and 18429 interactions. Surprisingly, this network included a core network with 1886 genes interacting with one another. This core network included 14 putative causal uveitis genes. Not included in the core network were 427 genes, of which 5 were uveitis genes, (*SELENOV*, *OR2L13*, *CAVIN4*, *DPP10*, and *HS3ST5*) none of which have specific expression or function associated with the eye or immune system.^33^

**Figure 5.**
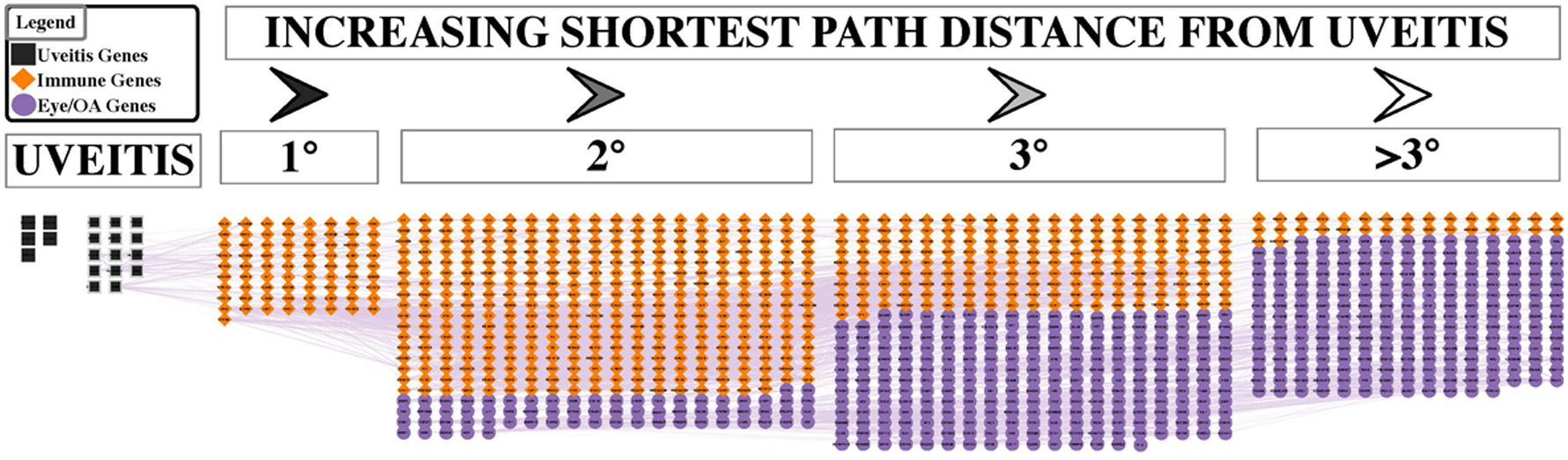
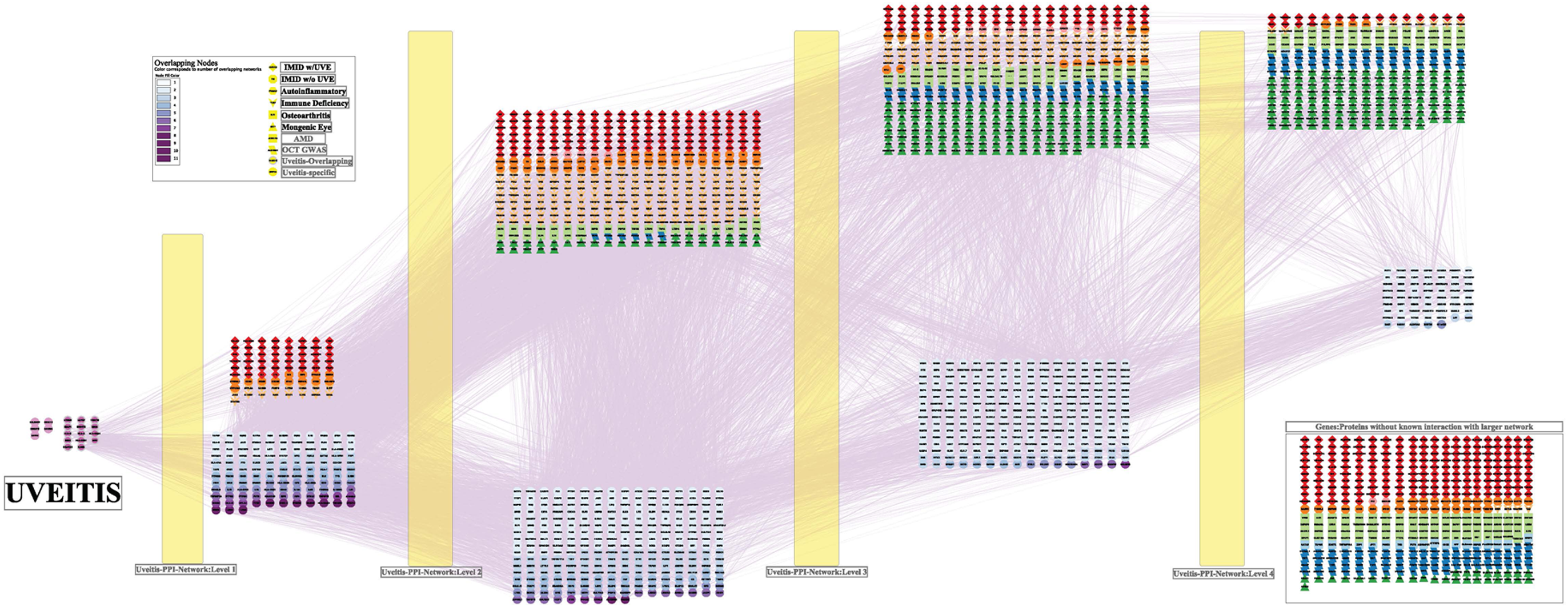
Uveitis is more closely related across biologic networks to immune-mediated diseases than to other ocular diseases. Uveitis genes are indicated as Black Squares. Immune genes are indicated as Orange Diamonds. Eye/Osteoarthritis (OA) genes are indicated as Purple Circles. Interaction edges are indicated by light purple lines. Genes are arranged in groups according to shortest path distance from the uveitis genes. Groups include first (1°), second (2°), third (3°) and fourth or more degree (>3°) neighbors of the Uveitis genes within the interaction network.

We found that the genes closest to uveitis genes in terms of geodesic or shortest-path distance from uveitis genes (Uveitis Genes) were predominately implicated in immune phenotypes (Immune Genes) (Figure 5). In contrast, genes included in ocular and osteoarthritis (for simplicity, denoted Eye Genes) were predominately those further away from the uveitis genes. To quantify our intuition, we also applied chi-squared analysis to the levels (degree of shortest path distance closeness) of this network. We found significant association with closeness to Uveitis Genes with Immune Genes (P < 4.5E-86). Strikingly, the genes one degree away from uveitis genes (first degree/1°) were 496 times more likely to be Immune Genes compared to those more than three degrees away (fourth degree or more/>3°) (Figure 5). Full network including genes implicated in multiple conditions is included in Supplemental Figure 1. Taken together, these findings support dysregulated inflammatory response as the primary genetic etiopathogenesis of non-infectious uveitis.

## 4. Discussion

Clinical association with IMIDs in approximately one-third of uveitis cases^10,11^ suggested a potential overlap in immune mechanisms. While prior narrative reviews genetics have highlighted several loci shared between uveitis and clinically-associated IMIDs,^34^ the genetic association between uveitis and IMIDs had not previously been systematically explored. Furthermore, since most uveitis cases are *not* associated with extraocular inflammation, the alternative hypothesis, that eye-specific factors drive ocular inflammation in uveitis, had also not been tested. Given this context, we sought to investigate the overlap in both putative causal genes and associated biologic networks between uveitis and both extraocular inflammatory diseases (IMIDs) and structural and degenerative eye diseases.

This computational synthesis of published GWAS data is consistent with the hypothesis that non-infectious uveitis shares significant genetic architecture with extraocular inflammatory disease, and not with other eye-related diseases. Both the high degree of causal gene overlap (75% with IMIDs and none with ocular diseases) and the relatively high protein-protein interaction network similarity support this hypothesis. While our analysis does to confirm shared disease mechanisms, it generates testable hypotheses that support future studies aimed at both expanded GWAS in uveitis and functional validation of putative shared disease mechanisms.

Autoimmune conditions cluster in families, between monozygotic twins and within individuals, revealing a strong genetic component to IMIDs that has been supported by GWAS evidence linking both specific variants and putative causal genes to multiple IMIDs, including HLA loci, *ERAP1/2*, *CTLA4*, *PTPN22*, *IL2RA*.^35,36^ To our knowledge, this computational analysis represents the first systematic exploration of pleiotropy between IMIDs and uveitis, and extends prior work associating uveitis with individual inflammatory diseases such as AS^1^. The HLA region, particularly HLA-DRB1, emerged as the most frequently shared locus between IMIDs and uveitis. This strongly reinforces prior findings that HLA genes drive a substantial proportion of the heritability and pleiotropy in autoimmunity. Their high polymorphism and central role in antigen presentation may explain why these loci repeatedly appear across diverse conditions. The high overlap of *HLA-DRB1*, *HLA-DQA1*, and *HLA-A/B*, as well as *ERAP1* between uveitis and diseases such as JIA, RA, MS, and UC suggests a core set of antigen presentation alleles may confer a pan-autoimmune risk, one that includes ocular inflammation. The genomic overlap between uveitis and AS is in line with recent cellular analyses. The presence of similar myeloid cell populations as well as CD8 T cells with shared TCRs in both ocular and joint infiltrates from patients with uveitis and arthritis genetically linked by HLA-B27^37,38^, supports a prominent role for antigen presentation pathways implicated by genetic overlap in MHC genes and *ERAP1*.

Importantly, non-antigen-presentation genes including *IL23R*, *MICA*, and *EGR2* also show significant overlap. These genes are involved in cytokine signaling (*IL23R*), immune cell activation (*MICA*) and transcriptional regulation (*EGR2*).^39–42^ Their involvement implicates immune processes beyond antigen presentation in the shared etiology of IMIDs and uveitis. Taken together, this analysis highlights two complementary axes of genetic convergence between uveitis and other diseases. This distinction offers a refined lens for interpreting disease mechanisms and may inform targeted therapeutic strategies that modulate either antigen presentation or common immune signaling pathways like IL-23.

Based on causal gene overlap and network similarity, uveitis was most like ankylosing spondylitis, sarcoidosis, Behcet’s disease, and JIA. This genetic similarity may relate to higher rates of clinical comorbidity, uveitis affects 16-48% of patients with Behcet’s disease^43–46^, 33-40% of patients with AS^8,27^, 5-30% of patients with sarcoidosis^28–31^, and 5.5-20% of patients with JIA^47–49^ compared to other IMIDs like SLE, in which the prevalence of uveitis is likely under 5%.^50,51^ The strong overlap between these IMIDs and uveitis supports the hypothesis that uveitis may share genetic pathways with other systemic inflammatory diseases, providing insight into shared disease mechanisms that could inform both diagnostic and therapeutic approaches. While uveitis shared 20% of causal genes with sarcoidosis, along with an extensive overlap of edges within the STRING protein-protein interaction network, the total number of cases in GWAS studies in either disease is low in comparison to GWAS of other immune diseases. Thus, genomic analyses of more uveitis cases are critical to better understand the genetic similarity between these two conditions.

### Study Limitations

Several limitations should be considered when interpreting our findings. First, the analysis was limited to a predefined set of inflammatory diseases, prioritizing those with available and well-powered GWAS data. This selection, while practical, may have excluded relevant conditions with emerging or underpowered genetic datasets, potentially biasing the observed overlap landscape. Additionally, uveitis itself is a heterogeneous condition encompassing multiple subtypes (e.g., anterior, intermediate, posterior), each of which may have distinct genetic drivers. Our analysis did not stratify by these subtypes due to limited subtype-specific summary statistics, which may have masked more nuanced associations. The genetic architecture of intermediate, posterior and panuveitis may differ from that of anterior uveitis and remain largely uncharacterized. There is an urgent need for subtype-stratified GWAS with deep clinical phenotyping to test whether IMID-associated endotypes map to discrete clinical uveitis phenotypes.

Methodologically, gene-level overlap was derived from curated loci reported in GWAS catalogues or high-confidence variant-to-gene mappings, which can vary in accuracy and resolution. By relying on lead variants/signals from multiple GWAS studies, we are at risk of double counting genes at individual loci. For example, the overlapping putative genes for uveitis and sarcoidosis consist of four predicted causal genes (*IL23R*, *C1orf141*, *HLA-A* and *HLA-DRB1*) at two physically separate genomic loci (chromosome 1: ∼67Mb and chromosome 6: ∼32Mb, positions hg38). However, the HLA associations with both sarcoidosis and anterior uveitis include multiple signals at the HLA locus.^52,53^ Additionally, there is evidence of complex association with multiple independent signals at *C1orf141-IL23R* in related immune diseases. In IBD, for example, traditional fine mapping and stepwise/adjusted logistic regression modeling identified multiple independent signals at *C1orf141-IL23R* locus.^54^ Likewise, publicly available summary statistics from the FinnGen-UKBiobank genome-wide PheWAS meta-analysis, the LD structure at this locus suggest the presence of multiple independent signals in sarcoidosis that may be shared with uveitis (which is labeled as iridocyclitis).^55,56^ Taken together, this example suggests that while our method may double count multiple genes at a particular locus, it may also *potentially* identify genomic regions with multiple signals at the same locus.

While overlap suggests shared genetic architecture, it does not confirm shared causality or mechanistic convergence without complementary functional or pathway analyses. Establishing mechanistic overlap will require additional studies in independent cohorts, including fine-mapping or colocalization and ultimately functional validation studies.

Biobank-based GWAS studies used in our computation synthesis were limited by the use of generic diagnostic codes (e.g. ICD10), rather than detailed clinical/deep phenotyping in the majority of the included GWAS studies. Future research should focus on in-depth phenotyping of uveitis and co-associated diseases, meta-analyses across inflammatory cohorts, and expanded GWAS efforts targeting uveitis subtypes, including anterior, intermediate, posterior, and panuveitis. These approaches will help disentangle shared versus subtype-specific genetic risk factors and refine our understanding of how inflammatory diseases intersect at the molecular level.

### Conclusion

This study provides a computational synthesis of the genetic architecture shared between uveitis and IMIDs and supports the hypothesis that uveitis shares pathophysiologic drivers with IMIDs such as AS, sarcoidosis, Crohn’s disease, and JIA. In contrast, minimal overlap was observed with structural or degenerative eye diseases, underscoring uveitis’ distinct immunogenetic profile.

This study reveals several testably hypotheses for future translational studies. 1) Fine mapping and colocalization studies of loci shared between uveitis and IMIDs will refine the identities of causal variants; 2) GWAS on an expanded cohort of phenotypically subtyped uveitis cases will reveal mechanistic disease endotypes that are share discrete immunogenetic architecture with systemic inflammatory conditions and reveal endotype-specific therapeutic targets; and 3) genomic stratification by pathway-related variants (i.e. IL-23) will improve outcomes in therapeutic trials of drugs repurposed from systemic IMIDs.

Future studies harnessing the insights of expert clinicians and multimodal imaging to more precisely define clinical phenotypes may increase the sensitivity of genomic analyses in uveitis and potentially yield endotype discrimination. Ultimately, such insights could inform precision risk prediction and catalyze the development of targeted, vision-preserving therapies for patients with uveitis and related conditions.

## Data Availability

all data are publically available, all code is provided via a link embedded in manuscript

https://github.com/chau-k/Genomic-network-analysis-links-uveitis-with-systemic-inflammatory-diseases

## Data and Code Availability Statement

The datasets generated and analyzed during the current study, along with all associated scripts used for processing and visualization, are publicly available on GitHub at https://github.com/chau-k/Genomic-network-analysis-links-uveitis-with-systemic-inflammatory-diseases.

## Author Contributions

LMH conceptualization, data curation, methodology, project administration, supervision, writing - original draft, writing- review and editing

KC Data curation, formal analysis, software, visualization, writing – original draft, writing – review and editing. KA writing- review and editing

TB writing- review and editing

ITWH, conceptualization, data curation, formal analysis, software, visualization, original draft, writing – review and editing.

## References

1. American Uveitis Society - Home. https://www.uveitissociety.org/.

2. Thorne, J. E. et al. Prevalence of Noninfectious Uveitis in the United States: A Claims-Based Analysis. JAMA Ophthalmol. 134, 1237–1245 (2016).

3. Gangaputra, S. S. et al. Effectiveness of Frequently Used TNF Inhibitors vs. Conventional Immunosuppressive Therapies for Noninfectious Uveitis. Ocul. Immunol. Inflamm. 33, 948–956 (2025).

4. Merrill, P. T. et al. Efficacy of Adalimumab in Non-Infectious Uveitis Across Different Etiologies: A Post Hoc Analysis of the VISUAL I and VISUAL II Trials. Ocul. Immunol. Inflamm. 29, 1569–1575 (2021).

5. Goto, H. et al. Adalimumab in Active and Inactive, Non-Infectious Uveitis: Global Results from the VISUAL I and VISUAL II Trials. Ocul. Immunol. Inflamm. 27, 40–50 (2019).

6. Nussenblatt, R. B., Mittal, K. K., Ryan, S., Green, W. R. & Maumenee, A. E. Birdshot retinochoroidopathy associated with HLA-A29 antigen and immune responsiveness to retinal S-antigen. Am. J. Ophthalmol. 94, 147–158 (1982).

7. Robinson, P. C. et al. Genetic dissection of acute anterior uveitis reveals similarities and differences in associations observed with ankylosing spondylitis. Arthritis Rheumatol. Hoboken NJ 67, 140–151 (2015).

8. Linssen, A. et al. The lifetime cumulative incidence of acute anterior uveitis in a normal population and its relation to ankylosing spondylitis and histocompatibility antigen HLA-B27. Invest. Ophthalmol. Vis. Sci. 32, 2568–2578 (1991).

9. Brewerton, D. A. The genetics of acute anterior uveitis. Trans. Ophthalmol. Soc. U. K. 104 (Pt 3), 248–249 (1985).

10. Mitkova-Hristova, V. T. et al. Epidemiology of Uveitis from a Tertiary Referral Hospital in Bulgaria over a 13-Year Period. Diagn. Basel Switz. 15, 828 (2025).

11. Barisani-Asenbauer, T. et al. Uveitis- a rare disease often associated with systemic diseases and infections- a systematic review of 2619 patients. Orphanet J. Rare Dis. 7, 57 (2012).

12. Garman, L. et al. Genome-Wide Association Study of Ocular Sarcoidosis Confirms HLA Associations and Implicates Barrier Function and Autoimmunity in African Americans. Ocul. Immunol. Inflamm. 29, 244–249 (2021).

13. Zekavat, S. M. et al. Phenome- and genome-wide analyses of retinal optical coherence tomography images identify links between ocular and systemic health. Sci. Transl. Med. 16, eadg4517 (2024).

14. Ghoussaini, M. et al. Open Targets Genetics: systematic identification of trait-associated genes using large-scale genetics and functional genomics. Nucleic Acids Res. 49, D1311–D1320 (2020).

15. Prioritising causal genes at GWAS loci (L2G) | Open Targets Genetics Documentation. https://genetics-docs.opentargets.org/our-approach/prioritising-causal-genes-at-gwas-loci-l2g (2022).

16. Assigning Variants to Genes (V2G) | Open Targets Genetics Documentation. https://genetics-docs.opentargets.org/our-approach/data-pipeline (2022).

17. Szklarczyk, D. et al. The STRING database in 2025: protein networks with directionality of regulation. Nucleic Acids Res. 53, D730–D737 (2025).

18. Shannon, P. et al. Cytoscape: A Software Environment for Integrated Models of Biomolecular Interaction Networks. Genome Res. 13, 2498–2504 (2003).

19. Holten, D. & Van Wijk, J. J. Force Directed Edge Bundling for Graph Visualization. Comput. Graph. Forum 28, 983–990 (2009).

20. von Mering, C. et al. STRING: known and predicted protein-protein associations, integrated and transferred across organisms. Nucleic Acids Res. 33, D433–437 (2005).

21. FAQ - STRING Help. https://string-db.org/help/faq/.

22. Getting started - STRING Help. https://string-db.org/help/getting_started/#network.

23. About - STRING functional protein association networks. https://string-db.org/cgi/about?footer_active_subpage=content.

24. Liao, K. P. et al. Specific association of type 1 diabetes mellitus with anti-cyclic citrullinated peptide-positive rheumatoid arthritis. Arthritis Rheum. 60, 653–660 (2009).

25. FinnGen provides genetic insights from a well-phenotyped isolated population - PubMed. https://pubmed.ncbi.nlm.nih.gov/36653562/.

26. Harley, I. T. W., Allison, K. & Scofield, R. H. Polygenic autoimmune disease risk alleles impacting B cell tolerance act in concert across shared molecular networks in mouse and in humans. Front. Immunol. 13, 953439 (2022).

27. Zeboulon, N., Dougados, M. & Gossec, L. Prevalence and characteristics of uveitis in the spondyloarthropathies: a systematic literature review. Ann. Rheum. Dis. 67, 955–959 (2008).

28. Ungprasert, P., Tooley, A. A., Crowson, C. S., Matteson, E. L. & Smith, W. M. Clinical Characteristics of Ocular Sarcoidosis: A Population-Based Study 1976-2013. Ocul. Immunol. Inflamm. 27, 389–395 (2019).

29. Birnbaum, A. D., French, D. D., Mirsaeidi, M. & Wehrli, S. Sarcoidosis in the national veteran population: association of ocular inflammation and mortality. Ophthalmology 122, 934–938 (2015).

30. Kuč, S. et al. Clinical Patterns of Sarcoidosis Patients with and without Uveitis: Insights from a Dutch Sarcoidosis Centre. Ocul. Immunol. Inflamm. 33, 125–132 (2025).

31. Shin, D., Kim, S., Kong, M., Kim, B. H. & Song, S. J. Prevalence and Incidence of Uveitis in Korean Patients with Sarcoidosis: A Population-Based Study. Ocul. Immunol. Inflamm. 33, 613–618 (2025).

32. Harley, I. T. W., Allison, K. & Scofield, R. H. Polygenic autoimmune disease risk alleles impacting B cell tolerance act in concert across shared molecular networks in mouse and in humans. Front. Immunol. 13, 953439 (2022).

33. The Human Protein Atlas. https://www.proteinatlas.org/.

34. Huang, X.-F. & Brown, M. A. Progress in the genetics of uveitis. Genes Immun. 23, 57–65 (2022).

35. Bogdanos, D. P. et al. Twin studies in autoimmune disease: genetics, gender and environment. J. Autoimmun. 38, J156–169 (2012).

36. Bao, Y. K. et al. High prevalence of comorbid autoimmune diseases in adults with type 1 diabetes from the HealthFacts database. J. Diabetes 11, 273–279 (2019).

37. Concepcion, C. et al. Compositional variation in eye-infiltrating immune cells distinguishes human uveitis subtypes. iScience 28, 111928 (2025).

38. Yang, X. et al. Autoimmunity-associated T cell receptors recognize HLA-B*27-bound peptides. Nature 612, 771–777 (2022).

39. McGovern, D. & Powrie, F. The IL23 axis plays a key role in the pathogenesis of IBD. Gut 56, 1333–1336 (2007).

40. Admon, A. ERAP1 shapes just part of the immunopeptidome. Hum. Immunol. 80, 296–301 (2019).

41. Zhu, W. et al. MICA*049, not MICA*009, is associated with Behçet’s disease in a Chinese population. Sci. Rep. 9, 10856 (2019).

42. Gómez-Martín, D., Díaz-Zamudio, M., Galindo-Campos, M. & Alcocer-Varela, J. Early growth response transcription factors and the modulation of immune response: Implications towards autoimmunity. Autoimmun. Rev. 9, 454–458 (2010).

43. Chadli, S. et al. Identification of clinical phenotypes in Behçet’s syndrome using latent class analysis: a step toward precision medicine. Clin. Exp. Rheumatol. 43, 1789–1798 (2025).

44. She, C.-H., Cai, J.-F., Hu, D., Bao, H.-F. & Guan, J.-L. Clinical characteristics of Behçet’s syndrome in Shanghai database: Baseline data of a cross-sectional cohort study. Int. J. Rheum. Dis. 27, e15355 (2024).

45. Nokhatha, S. A. A. et al. High prevalence of HLA-B51 and ocular involvement in Behçet’s disease: a multicenter cross-sectional study. Rheumatol. Int. 45, 140 (2025).

46. Lee, B. H., Chung, K. B., Jang, H., Jung, Y. W. & Kim, D.-Y. HLA-B51 Positivity Correlates With Symptom Completeness From Recurrent Aphthous Stomatitis to Complete Behçet’s Disease. J. Dermatol. 52, 1001–1007 (2025).

47. Grassi, A., Corona, F., Casellato, A., Carnelli, V. & Bardare, M. Prevalence and outcome of juvenile idiopathic arthritis-associated uveitis and relation to articular disease. J. Rheumatol. 34, 1139–1145 (2007).

48. Heiligenhaus, A. et al. Prevalence and complications of uveitis in juvenile idiopathic arthritis in a population-based nation-wide study in Germany: suggested modification of the current screening guidelines. Rheumatol. Oxf. Engl. 46, 1015–1019 (2007).

49. Marshall, R., Thorne, J. & Berkenstock, M. K. Prevalence and incidence of juvenile idiopathic arthritis-associated uveitis and medication trends in the TriNetX database. Br. J. Ophthalmol. bjo-2024-327087 (2025) doi:10.1136/bjo-2024-327087.

50. Teng, R. W., Estrela, T., Meidan, E. & Gise, R. Ophthalmic manifestations of systemic lupus erythematosus at a quaternary children’s hospital. J. AAPOS Off. Publ. Am. Assoc. Pediatr. Ophthalmol. Strabismus 29, 104276 (2025).

51. Cheng, T. et al. Clinical features of ocular damage in systemic lupus erythematosus and risk factors for hydroxychloroquine-related complications. Surv. Ophthalmol. 69, 733–742 (2024).

52. Rybicki, B. A. & Iannuzzi, M. C. Sarcoidosis and human leukocyte antigen class I and II genes: it takes two to tango? Am. J. Respir. Crit. Care Med. 169, 665–666 (2004).

53. Gelfman, S. et al. A large meta-analysis identifies genes associated with anterior uveitis. Nat. Commun. 14, 7300 (2023).

54. Rivas, M. A. et al. Deep resequencing of GWAS loci identifies independent rare variants associated with inflammatory bowel disease. Nat. Genet. 43, 1066–1073 (2011).

55. gene IL23R H7_IRIDOCYCLITIS. https://public-metaresults-fg-ukbb.finngen.fi/gene/IL23R/pheno/H7_IRIDOCYCLITIS.

56. gene IL23R D3_SARCOIDOSIS. https://public-metaresults-fg-ukbb.finngen.fi/gene/IL23R/pheno/D3_SARCOIDOSIS.

