## Supplementary figures and images for "Genomic network analysis links uveitis with systemic inflammatory diseases" (from another version of this preprint: version 1, 2026-03-26)

### Figure S1 Predicted causal risk genes for uveitis are closer* to inflammatory diseases than structural eye disease.

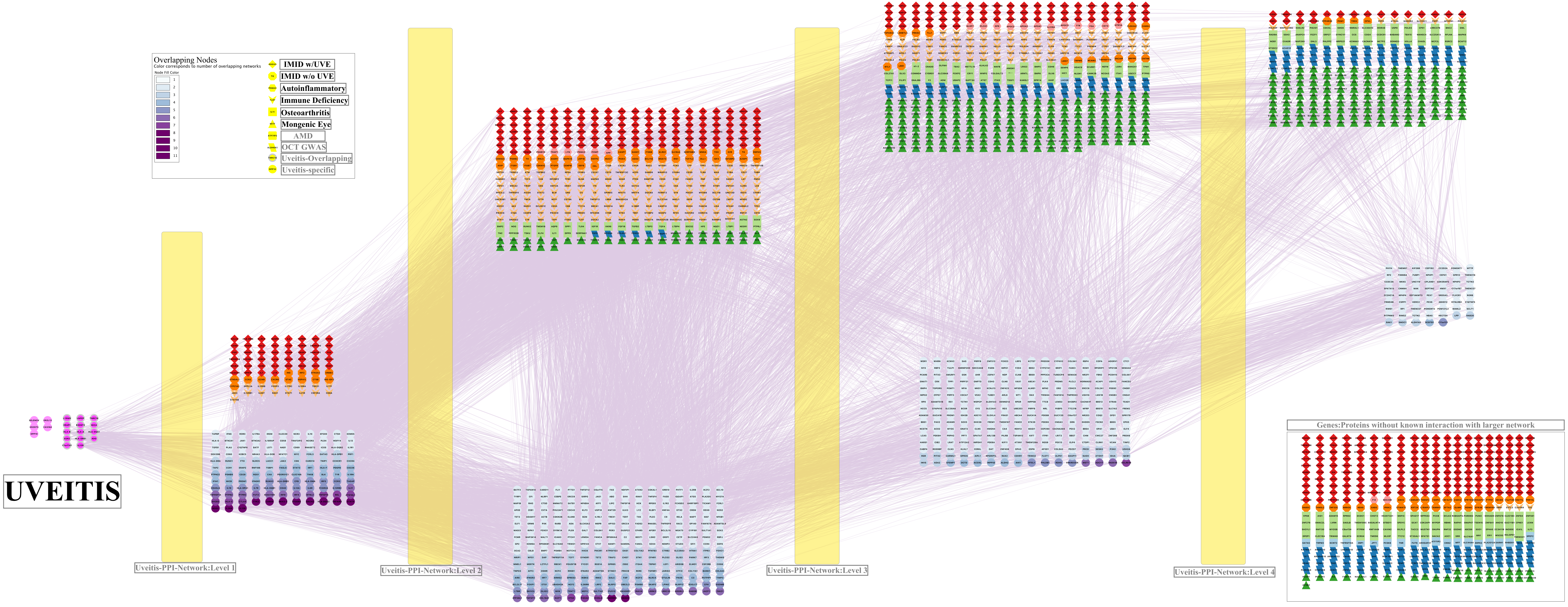
